# Comparison of the carotid corrected flow time and tidal volume challenge for assessing fluid responsiveness in robot-assisted laparoscopic surgery

**DOI:** 10.1101/2023.04.10.23288384

**Authors:** Xixi Tang, Qi Chen, Zejun Huang, Ran An, Jingqiu Liang, Hongliang Liu

## Abstract

**Purpose:** We aimed to evaluate the ability of carotid corrected flow time assessed by ultrasound to predict fluid responsiveness in patients undergoing robot-assisted laparoscopic gynecological surgery in the modified head-down lithotomy position.

**Methods:** This prospective single-center study conducted at the Chongqing University Cancer Hospital included patients undergoing robot-assisted laparoscopic surgery in the modified head-down lithotomy position. Carotid doppler parameters and hemodynamic data, including corrected flow time, pulse pressure variation (PPV), stroke volume variation, and stroke volume index at a tidal volume of 6 mL/kg predicted body weight and after increasing the tidal volume to 8 mL/kg predicted body weight (tidal volume challenge), respectively, were measured. Fluid responsiveness was defined as a stroke volume index ≥10% increase after volume expansion.

**Results:** Among the 52 patients included, 26 were classified as fluid responders and 26 as non-responders based on the stroke volume index. The area under the receiver operating characteristic curve values measured to predict the fluid responsiveness to corrected flow time and changes in PPV (ΔPPV_6–8_) after tidal volume challenge were 0.82 [95% confidence interval (CI): 0.705–0.937; P < 0.0001] and 0.85 (95% CI: 0.740–0.956; P < 0.0001), respectively. Both values were higher than those for PPV at a tidal volume of 8 mL/kg (0.79, 95% CI: 0.674–0.911; P = 0003). The optimal cut-off values for corrected flow time and ΔPPV_6–8_ were 356.5 ms and >1%, respectively.

**Conclusion:** The change in PPV after tidal volume challenge and corrected flow time reliably predicted fluid responsiveness in patients undergoing robot-assisted laparoscopic gynecological surgery in the modified head-down lithotomy position.

**Trial registration:** Chinese Clinical Trial Register (CHiCTR2200060573)

## Introduction

The choice between “liberal” and “restrictive” fluid management remains controversial. Preoperative fasting, bowel preparation, and third space loss can lead to hypovolemia. Moreover, inadequate intraoperative administration of fluids can result in poor outcomes, including acute tubular necrosis. However, fluid overload increases the burden on the heart and pulmonary congestion, decreases tissue oxygenation, inhibits wound healing, and delays recovery. Therefore, appropriate perioperative infusion management is essential for optimizing perioperative outcomes [1]. Patients with increased stroke volume (SV) after adequate fluid resuscitation are considered “fluid responsive.”

Robot-assisted surgery is used extensively owing to its advantages of smaller surgical wounds, early recovery, clearer vision, and manipulation dexterity [2]. Venous return increases cardiac output through the modified head-down lithotomy positioning; however, further increases in intra-abdominal pressure decrease cardiac output. Compression of the abdominal aorta, production of neurohumoral factors, and activation of the renin-angiotensin-aldosterone axis increase systemic vascular resistance and depress myocardial contractility [3], thus complicating fluid management.

Dynamic changes, pulse pressure variation (PPV) and stroke volume variation (SVV), in arterial waveform-derived variables, are superior to traditional static indicators such as central venous pressure and pulmonary artery occlusion pressure [4–6]. PPV and SVV are based on heart-lung interactions, and their reliability is incompletely proven during spontaneous breathing, arrhythmias, low tidal volume (VT), elevated intra-abdominal pressure, high respiratory rate, and right heart failure [4]. The absolute changes in PPV and SVV values induced by VT challenge predict fluid responsiveness with high sensitivity and specificity; however, VT challenge may not overcome the other limitations associated with PPV and SVV use while a patient is spontaneously breathing or has increased intra-abdominal pressure [6,7]. Furthermore, hemodynamic monitoring techniques are often invasive and expensive and can lead to infection, hematoma, peripheral ischemia, nerve injury, and perforation [8,9].

Carotid ultrasound measurement techniques have the advantages of simple operation and non-invasive, easy-to-obtain, and straightforward display of the measured data. There is a direct and significant correlation between carotid corrected flow time (FTc) and intravascular volume status [10]. Carotid FTc is a reliable predictor of fluid responsiveness during spontaneous breathing or mechanical ventilation [11,12].

This study investigated the feasibility of using carotid FTc measured via Doppler ultrasound to predict fluid responsiveness in patients undergoing robot-assisted laparoscopic surgery in the modified head-down lithotomy position. Further, we compared the ability of carotid FTc with those of the changes in PPV and SVV observed after VT challenge to predict fluid responsiveness.

## Methods

### Study population

This study was approved by the Institutional Review Board of Chongqing University Cancer Hospital (approval number: CZLS2021041-A) and registered before patient enrollment on the Chinese Clinical Trial Register (CHiCTR2200060573). Fifty-five patients with an American Society of Anesthesiologists physical status of class I−III, scheduled to undergo robot-assisted laparoscopic surgery with pneumoperitoneum in the modified head-down lithotomy position, were enrolled after obtaining written informed consent(Fig 1). The exclusion criteria were body mass index of >30 or <15 kg/m^2^; arrhythmia; moderate-to-severe valvular heart disease; presence of >50% carotid artery stenosis by conventional angiography, computed tomographic angiography, magnetic resonance angiography, duplex ultrasonography, or newly detected carotid artery stenosis during the study period; left ventricular ejection fraction of <50%; right ventricular dysfunction; moderate-to-severe chronic obstructive pulmonary disease; chronic kidney disease; and pregnancy.

**Figure 1.**
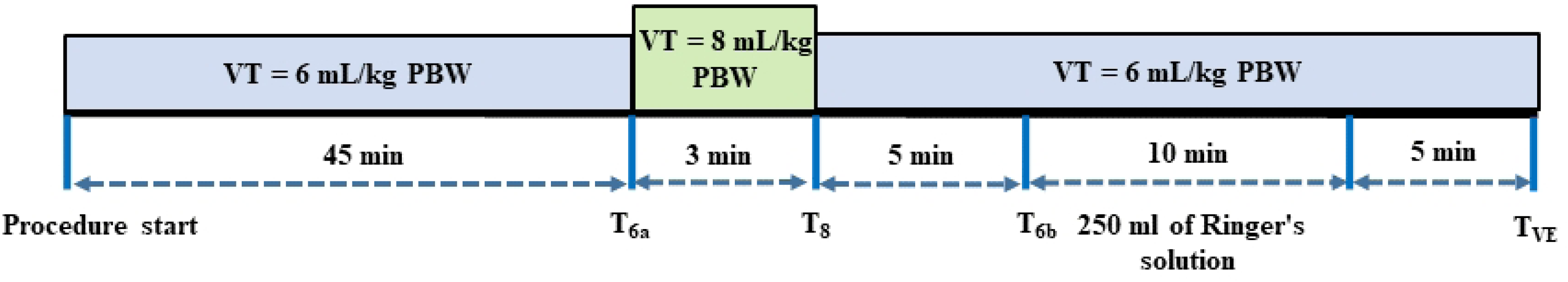
Study design. T_6a_: Time at a tidal volume of 6 mL/kg predicted body weight; T_8_: Time at a tidal volume of 8 mL/kg predicted body weight; T_6b_: Time at a tidal volume of 6 mL/kg predicted body weight; T_VE_: Time after volume expansion; VT: Tidal volume; PBW: Predicted body weight

### Anesthesia technique

After the patients entered the operating room, the following parameters were monitored using standard intraoperative methods: heart rate (HR), peripheral oxygen saturation, continuous electrocardiographic values, and non-invasive blood pressure. Anesthesia was induced with midazolam (1−2 mg), propofol (2−3 mg/kg), sufentanil (0.3−0.5 µg/kg), and rocuronium (0.6−0.9 mg/kg). Anesthesia was maintained with a continuous infusion of propofol (1.5−3 mg/kg/h), sevoflurane (1−3 vol%), and remifentanil (0.02−0.2 μg/kg/min). Neuromuscular blockade was maintained throughout each procedure via the intermittent administration of rocuronium bromide (0.15 mg/kg every 30−40 min).

After tracheal intubation, VT was maintained at 6 mL/kg of the predicted body weight (PBW) (determined as x + 0.91[height (in cm) - 152.4], where x = 50 for males and 45.5 for females) [13], and positive end-expiratory pressure was set at 5 cmH_2_O. An end-tidal carbon dioxide concentration of 35−45 mmHg was maintained by adjusting the ventilation frequency. All respiratory parameters, including plateau pressure (Pplat) and compliance of the respiratory system (Crs), were recorded using the WATO EX-65 anesthesia machine (Mindray Medical Systems, Shenzhen, China). All patients were placed in the modified head-down lithotomy position during the surgery. Pneumoperitoneum was maintained via continuous carbon dioxide insufflation to maintain an intra-abdominal pressure of 12 mmHg, and 4 mL/kg/h Ringer’s solution was administered for fluid maintenance.

### Hemodynamic monitoring

A radial arterial catheter was inserted after anesthesia induction. The arterial pressure transducer was levelled and zeroed at the intersection of the anterior axillary line and fifth intercostal space. The arterial pressure signal was connected to an IntelliVue MP40 monitor (Philips Medizin Systeme Böblingen GmbH, Böblingen, Germany) and the MostCare device (Vygon, Vytech, Padova, Italy) using a Y cable. The square-wave test was performed to exclude under- or overdamping of the pressure signal [14–16].

MostCare, which has a sampling rate of 1.000 point per second, calculates SV using the following equation: SV = Asys/Ztot, where Asys is the area under the systolic part of the arterial pressure waveform and Ztot is systemic vascular resistance. SVV was calculated by assessing the changes in SV as follows: SVV = (maximum SV – minimum SV)/mean SV × 100. All indexed values, including stroke volume index (SVI), were calculated using the anthropometric measurements of each patient. Hemodynamic variables were recorded using MostCare according to the manufacturer’s default time setting (30 s) and imported into a dedicated Excel spreadsheet for further analysis [16]. The Philips monitor directly measured systolic and diastolic blood pressure and mean arterial pressure (MAP) from the arterial pressure waveform. The PPV value was calculated automatically and continuously recorded in real-time by the monitor. The pulse pressure (PP) was defined as the difference between the diastolic and systolic arterial pressure, and PPV was calculated as follows: PPV = (maximum PP − minimum PP)/mean PP × 100. All values were averages of the three consecutive measurements acquired.

### Carotid ultrasonography

Carotid ultrasound images were obtained using a portable ultrasound device (Mindray Medical Systems, Shenzhen, China) with the participants in the modified head-down lithotomy position. FTc was measured as previously described by Blehar et al [17]. A linear array (12 MHz) transducer was placed longitudinally on the neck with the probe marker pointing to the patient’s head. A long-axis B-mode image of the right common carotid artery was obtained at the level of the lower border of the thyroid cartilage. Spectral Doppler tracings were then obtained by placing a 0.5-mm sample gate through the center of the vessel, within 2−3 cm proximal to the carotid bulb in the longitudinal plane and an insonation angle controlled at ≤60° to accurately measure blood flow velocity [18,19]. After the pulsed-wave Doppler spectrum was displayed, the optimal sampling volume and angle were adjusted to obtain a satisfactory spectrum, and the image was frozen. Flow time (FT) was gauged from the beginning of the systolic upstroke to the dicrotic notch. FTc was calculated using Wodey’s formula as follows: FTc = FT + [1.29 × (HR – 60)] [20]. The examiner obtained FTc measurements during three continuous heartbeat periods, and all three values were averaged and recorded for subsequent analysis. HR was obtained by measuring the interval between the heartbeats at the beginning of the Doppler flow upstroke through two consecutive cycles (Fig 2).

**Figure 2.**
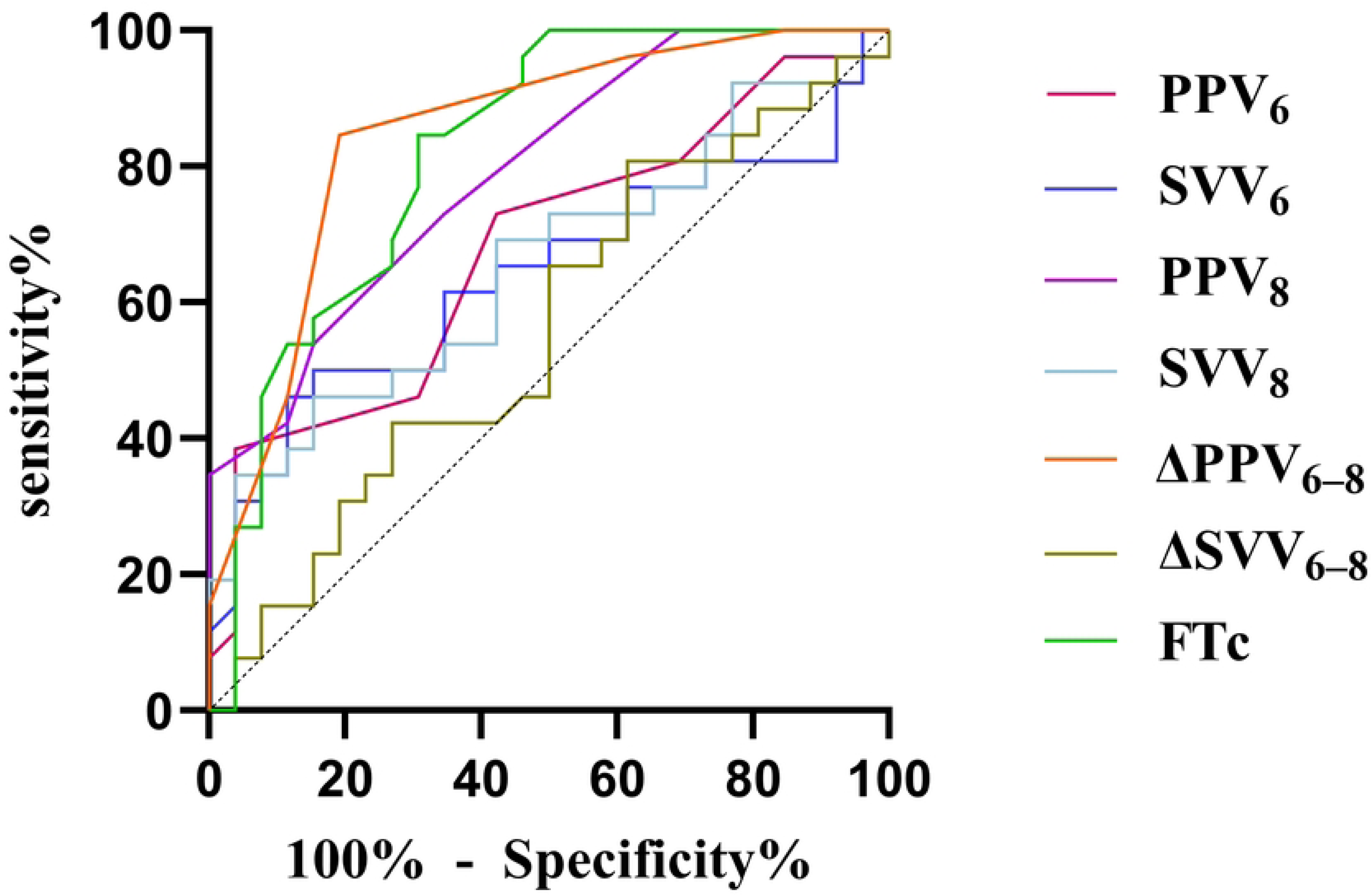
Comparison of the receiver operating characteristic curves in predicting the fluid responsiveness. PPV6: pulse pressure variation during tidal volume (VT) of 6 mL/kg predicted body weight (PBW); SVV6: stroke volume variation during VT of 6 mL/kg PBW; PPV8: pulse pressure variation during VT of 8 mL/kg PBW; SVV8: stroke volume variation during VT of 8 mL/kg PBW; ΔPPV_6–8_: change in the value of pulse pressure variation after VT challenge; ΔSVV_6–8_: change in the value of stroke volume variation after VT challenge; FTc: carotid corrected flow time of the artery

### Study procedures

Fig 3 illustrates the study design. No vasoactive medications were used during the measurement period, and all measurements were obtained during surgery while the patient was hemodynamically stable, defined as MAP and HR changes of <10% over 5 min. The protocol was initiated at least 45 min after increasing the intra-abdominal pressure to 12 mmHg, and the first set of measurements (HR, MAP, SVV_6_, PPV_6_, SVI, VT, Pplat, Crs, and FTc) was recorded concurrently (time-point T_6a_). After obtaining the baseline measurements, the VT setting was increased from 6 to 8 mL/kg PBW for 3 min (VT challenge), and the hemodynamic and respiratory variables, including SVV_8_ and PPV_8_, were measured again during the last minute at 8 mL/kg PBW VT ventilation (time-point T_8_). VT was then returned to 6 mL/kg PBW, and the third set of data was recorded after 5 min (time-point T_6b_). Subsequently, 250 mL of Ringer’s solution was injected over 10 min. The last set of parameters (time-point T_VE_) was recorded 5 min after fluid administration. The changes in PPV and SVV due to the VT challenge were calculated as follows: ΔPPV_6-8_ = PPV_8_ − PPV_6_; and ΔSVV_6-8_ = SVV_8_ − SVV_6_. Fluid responsiveness was defined as an SVI increase of ≥10% and assessed using the MostCare monitor after administering balanced crystalloids [21,22].

**Figure 3.**
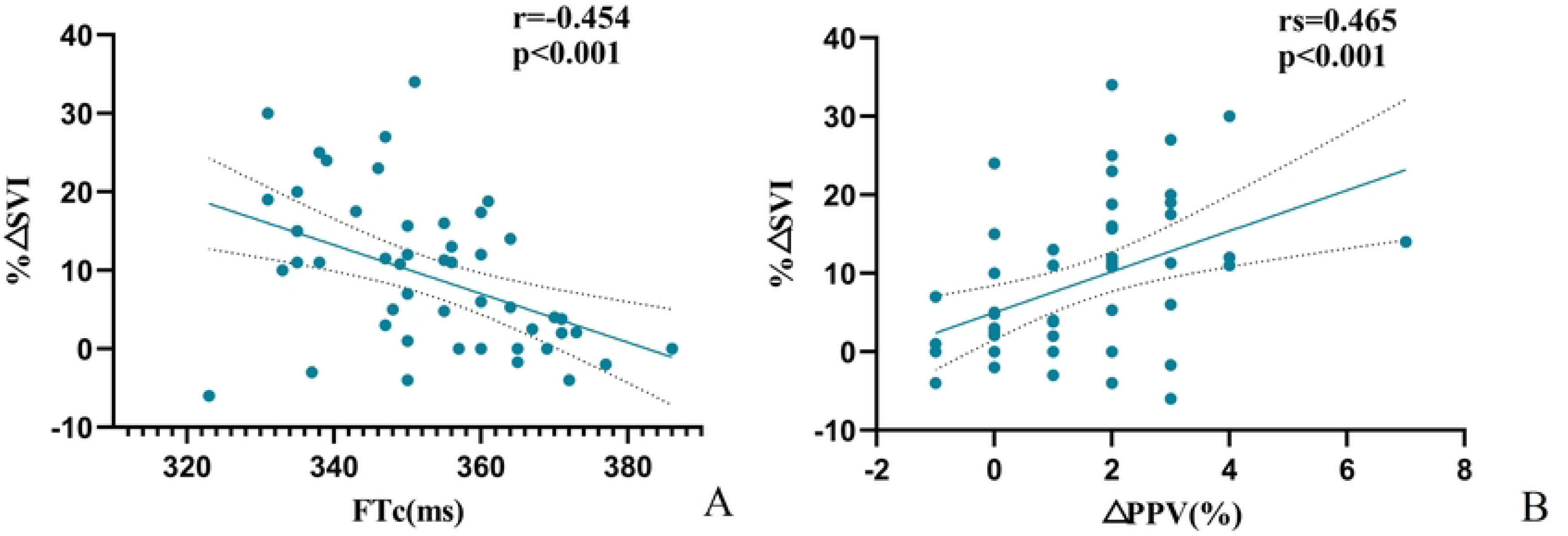
Relationship between the carotid corrected flow time (FTc) and the change in the value of pulse pressure variation after tidal volume challenge (ΔPPV_6–8_) and volume expansion-induced percentage changes in the stroke volume index (SVI) A: Relationship between FTc and SVI B: Relationship between ΔPPV_6–8_ and SVI

### Sample size calculation and statistical analysis

The sample size was calculated using tests for a one-curve module using PASS ver.15.0 (IBM Corp., Armonk, NY, USA). A previous study has reported that the area under the curve (AUC) for predicting fluid responsiveness using descending aorta FTc is 0.82 [23]. Therefore, we assumed a relatively low AUC of 0.75 for carotid FTc. At least 50 patients were required to compare this value to the null hypothesis (AUC = 0.50), with a type I error of 0.05, power of 0.90, and sample size of negative/positive group ratio of 1. A 10% dropout or withdrawal rate was estimated; therefore, enrollment of 55 patients was planned.

Distribution normality was assessed using the Kolmogorov−Smirnov test. Data are presented as median (interquartile range), mean (standard deviation), and the number of patients (%). The characteristics of responders and non-responders were compared using the independent t-test for non-normally distributed data and Mann−Whitney U test for non-normally distributed data, while categorical variables were analyzed using the chi-square test. Hemodynamic variables at T_6a_, T_8_, T6_b_, and T_VE_ were compared using the paired t-test or Wilcoxon signed-rank sum test. Between-group comparisons were performed using the t-test or Mann−Whitney U test.

A receiver operating characteristic curve analysis was performed to assess reliability, and the AUC values were compared using the DeLong test [24]. The best cut-off value was determined by maximizing the Youden index (sensitivity + specificity - 1) [25]. The cut-off values delimiting the grey zone were defined by the values associated with a 90% sensitivity and 90% specificity [11].

Relationships between the percentage change in SVI after VE and ΔPPV_6–8_ were assessed using the Spearman rank correlation test. Relationships between the percentage change in SVI after VE and carotid ultrasound FTc were assessed using Pearson correlation analysis.

Statistical analyses were performed using MedCalc ver. 20.1.0 (MedCalc Software, Ostend, Belgium), GraphPad Prism ver. 9.4.0 (GraphPad Software, San Diego, CA, USA), and SPSS ver. 27.0 (IBM Corp., Armonk, NY, USA). Statistical significance was set at P <0.05.

## Results

Among the 59 patients assessed for eligibility in the study, 55 were enrolled. Among them, three were excluded for the following reasons: high airway pressure (n = 1), severe hypotension (n = 1), and frequent intraoperative premature ventricular contraction (n = 1). Therefore, 52 patients were included in the final analysis (Fig 1). A between-group comparison of responders and non-responders revealed that patients’ clinical characteristics did not differ (Table 1).

**Table 1.** Patient characteristics

|  | Overall(n=52) | Responder(n=26) | Non-Responders(n=26) | P-Value |
| --- | --- | --- | --- | --- |
| Age, year | 50.98±10.9 | 51.4±10.1 | 50.5±11.8 | 0.773<sup>a</sup> |
| BMI, kg/m<sup>2</sup> | 23.6±3.2 | 23.6±3.6 | 23.7±2.8 | 0.966<sup>a</sup> |
| ASA physical status (II/III) | 43,9 | 22,4 | 21,5 | 1<sup>b</sup> |
| Diagnosis, n (%) |  |  |  | 0.637<sup>c</sup> |
| Vaginal cancer | 6(11.5) | 4(15.4) | 2(7.7) |  |
| Cervical Cancer | 28(53.8) | 14(53.8) | 14(53.8) |  |
| Endometrial cancer | 18(34.6) | 8(30.8) | 10(38.5) |  |
| Comorbidities, n (%) |  |  |  | 0.206<sup>c</sup> |
| Hypertension | 5(9.6) | 1(3.8) | 4(13.8) |  |
| Others | 5(9.6) | 4(13.8) | 1(3.8) |  |

Table 2 presents the hemodynamic and respiratory characteristics of the patients at each time point. The MAP values of responders were significantly higher than those of non-responders at all time points. In responders, when VT was increased to 8 mL/kg PBW, PPV and SVV significantly increased, and decreased significantly after VE. All responders had significantly lower SVI and FTc values than non-responders before VE; however, SVI and FTc increased in all patients after rehydration. Despite rehydration, the FTc values of the groups remained significantly different.

**Table 2.**
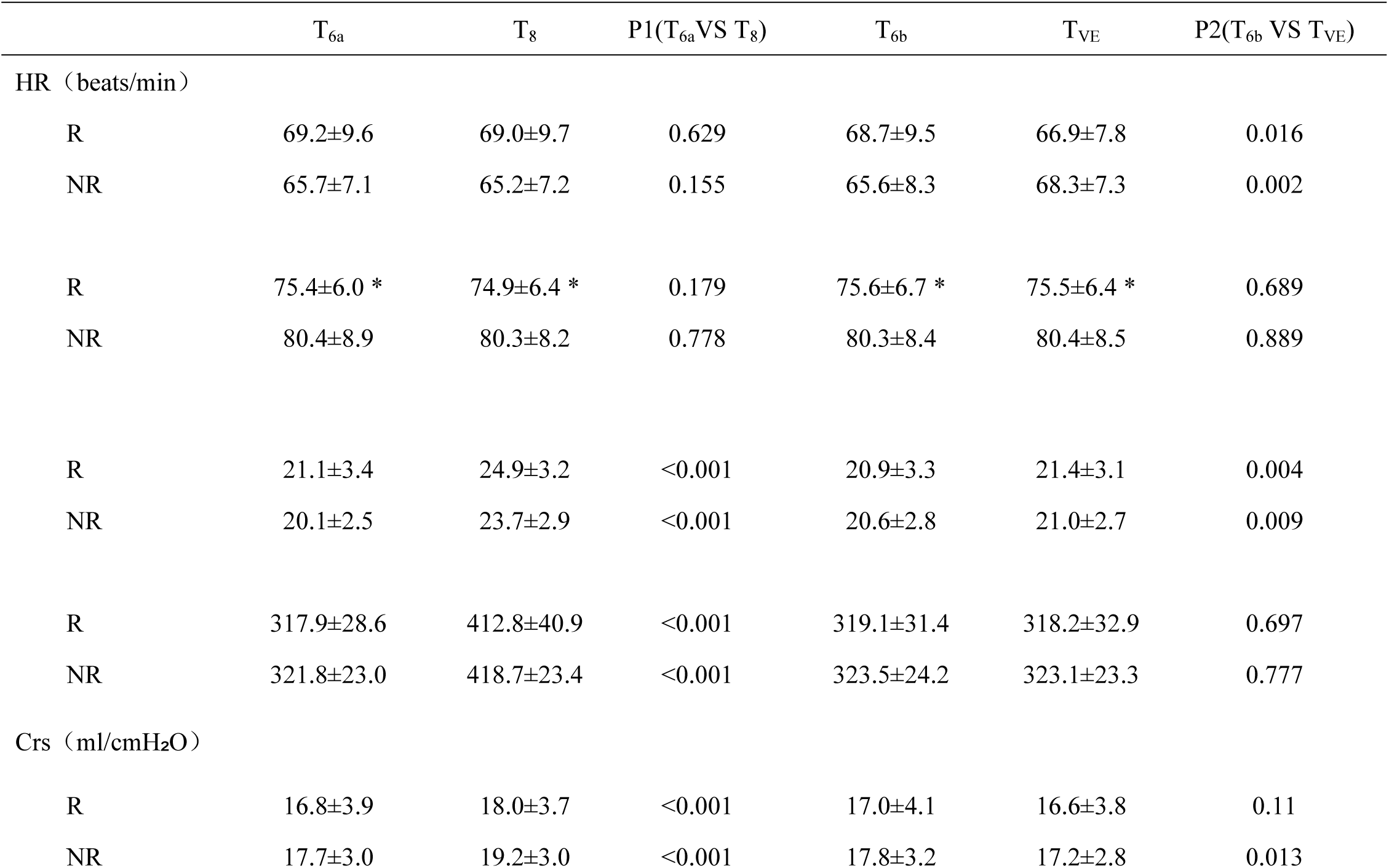
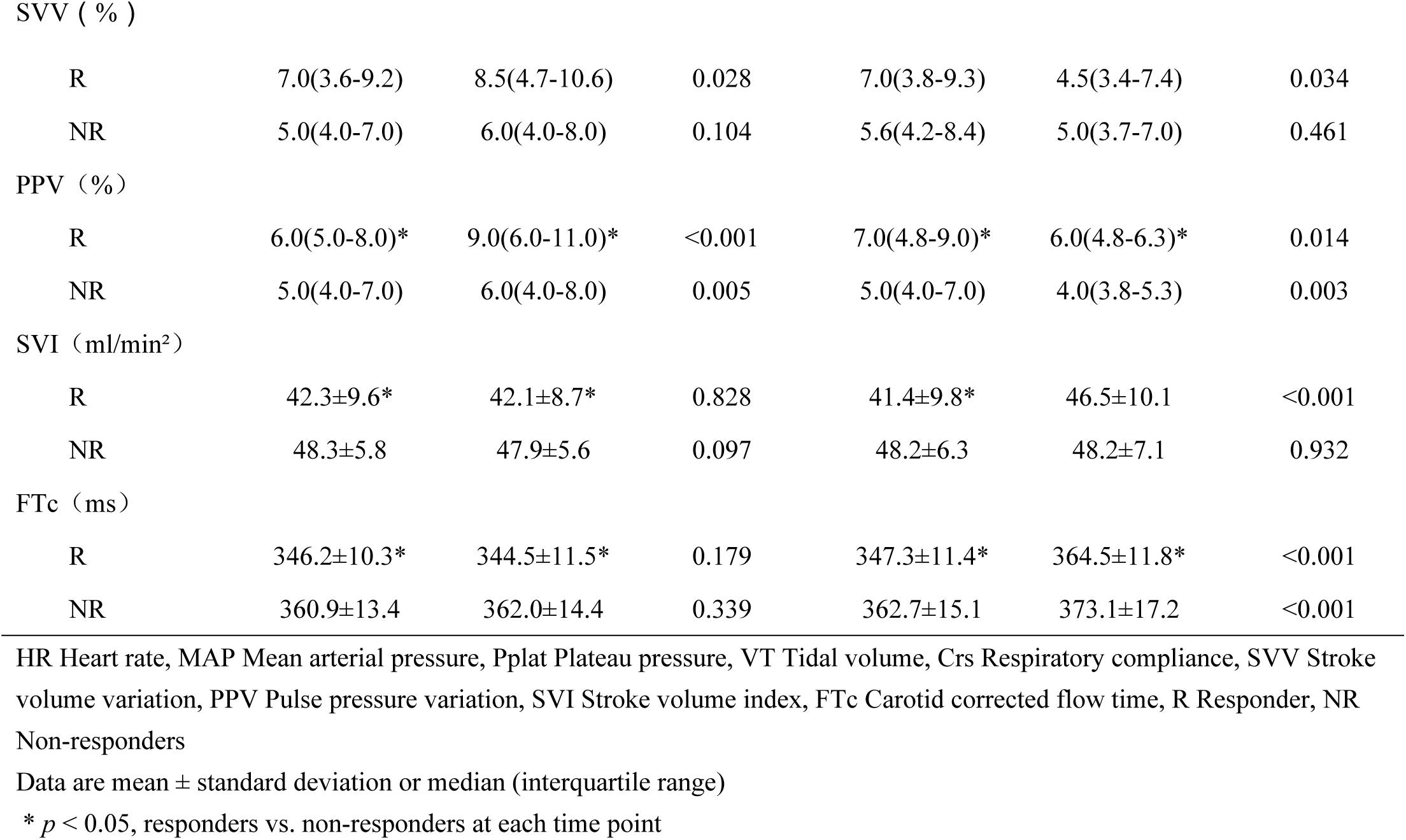
The hemodynamic and respiratory characteristics at baseline, after tidal ventilation challenge, before volume expansion, and after volume expansion

|  | T<sub>6a</sub> | T<sub>8</sub> | P1(T<sub>6a</sub> VS T<sub>8</sub>) | T<sub>6b</sub> | T<sub>VE</sub> | P2(T<sub>6b</sub> VS T<sub>VE</sub>) |
| --- | --- | --- | --- | --- | --- | --- |
| HR (beats/min) |  |  |  |  |  |  |
| R | 69.2±9.6 | 69.0±9.7 | 0.629 | 68.7±9.5 | 66.9±7.8 | 0.016 |
| NR | 65.7±7.1 | 65.2±7.2 | 0.155 | 65.6±8.3 | 68.3±7.3 | 0.002 |
| Map (mm Hg) |  |  |  |  |  |  |
| R | 75.4±6.0 * | 74.9±6.4 * | 0.179 | 75.6±6.7 * | 75.5±6.4 * | 0.689 |
| NR | 80.4±8.9 | 80.3±8.2 | 0.778 | 80.3±8.4 | 80.4±8.5 | 0.889 |
| Pplat (cm H<sub>2</sub>O) |  |  |  |  |  |  |
| R | 21.1±3.4 | 24.9±3.2 | <0.001 | 20.9±3.3 | 21.4±3.1 | 0.004 |
| NR | 20.1±2.5 | 23.7±2.9 | <0.001 | 20.6±2.8 | 21.0±2.7 | 0.009 |
| VT (ml) |  |  |  |  |  |  |
| R | 317.9±28.6 | 412.8±40.9 | <0.001 | 319.1±31.4 | 318.2±32.9 | 0.697 |
| NR | 321.8±23.0 | 418.7±23.4 | <0.001 | 323.5±24.2 | 323.1±23.3 | 0.777 |
| Crs (ml/cmH<sub>2</sub>O) |  |  |  |  |  |  |
| R | 16.8±3.9 | 18.0±3.7 | <0.001 | 17.0±4.1 | 16.6±3.8 | 0.11 |
| NR | 17.7±3.0 | 19.2±3.0 | <0.001 | 17.8±3.2 | 17.2±2.8 | 0.013 |
| SVV ( % ) |  |  |  |  |  |  |
| R | 7.0(3.6-9.2) | 8.5(4.7-10.6) | 0.028 | 7.0(3.8-9.3) | 4.5(3.4-7.4) | 0.034 |
| NR | 5.0(4.0-7.0) | 6.0(4.0-8.0) | 0.104 | 5.6(4.2-8.4) | 5.0(3.7-7.0) | 0.461 |
| PPV ( % ) |  |  |  |  |  |  |
| R | 6.0(5.0-8.0)* | 9.0(6.0-11.0)* | <0.001 | 7.0(4.8-9.0)* | 6.0(4.8-6.3)* | 0.014 |
| NR | 5.0(4.0-7.0) | 6.0(4.0-8.0) | 0.005 | 5.0(4.0-7.0) | 4.0(3.8-5.3) | 0.003 |
| SVI ( ml/min<sup>2</sup> ) |  |  |  |  |  |  |
| R | 42.3±9.6* | 42.1±8.7* | 0.828 | 41.4±9.8* | 46.5±10.1 | <0.001 |
| NR | 48.3±5.8 | 47.9±5.6 | 0.097 | 48.2±6.3 | 48.2±7.1 | 0.932 |
| FTc ( ms ) |  |  |  |  |  |  |
| R | 346.2±10.3* | 344.5±11.5* | 0.179 | 347.3±11.4* | 364.5±11.8* | <0.001 |
| NR | 360.9±13.4 | 362.0±14.4 | 0.339 | 362.7±15.1 | 373.1±17.2 | <0.001 |

HR Heart rate, MAP Mean arterial pressure, Pplat Plateau pressure, VT Tidal volume, Crs Respiratory compliance, SVV Stroke volume variation, PPV Pulse pressure variation, SVI Stroke volume index, FTc Carotid corrected flow time, R Responder, NR Non-responders
Data are mean ± standard deviation or median (interquartile range)
\* $p < 0.05$ , responders vs. non-responders at each time point

The AUC values of FTc and ΔPPV_6–8_ were 0.82 [95% confidence interval (CI): 0.705−0.937; P < 0.0001] and 0.85 (95% CI: 0.740−0.956; P < 0.0001), respectively, indicating both are excellent predictors of fluid responsiveness (Fig 4). The optimal cut-off values for FTc and ΔPPV_6–8_ were 356.5 ms (sensitivity, 84.6%; specificity, 69.2%) and >1% (sensitivity, 80.8%; specificity, 76.9%), respectively. The grey zones for FTc and ΔPPV_6–8_ were 347.1−359.9 ms and 0.3−2.7, respectively, containing 27% and 48% of patients, respectively (Table 3).

**Table 3.**
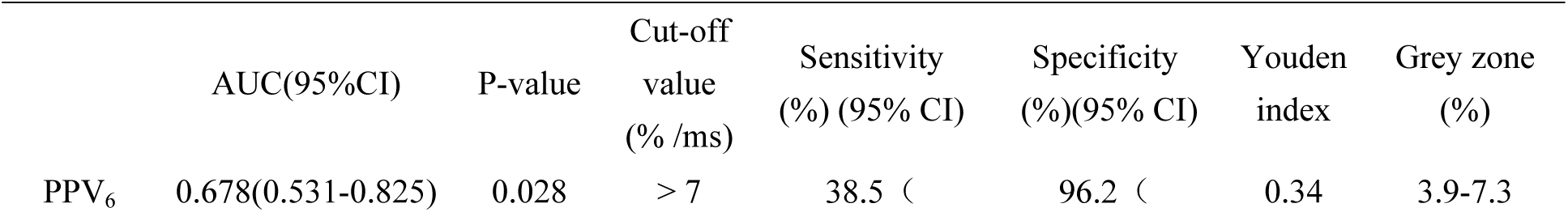
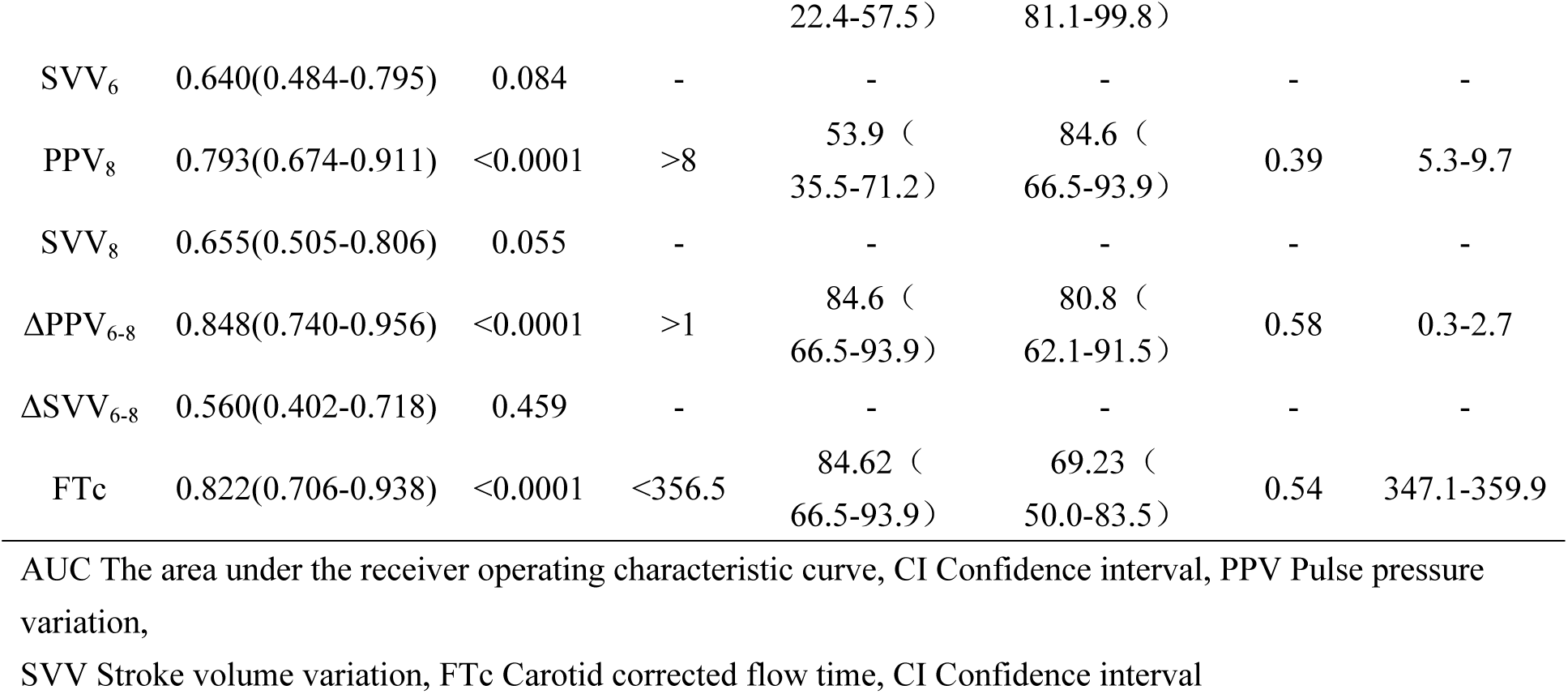
Diagnostic accuracy of various variables to predict fluid responsiveness

The predictive power of PPV_8_, with an AUC of 0.79 (95% CI: 0.674−0.911; P = 0.003), showed only fair capability. However, PPV_6_ was a poor predictor, with an AUC of only 0.68 (95% CI: 0.530−0.825; P = 0.028). Additionally, the SVV-related index did not significantly predict fluid responsiveness. The AUC values of ΔPPV_6–8_, FTc, and PPV_8_ did not differ significantly (P > 0.05).

FTc and ΔPPV_6–8_ were correlated with the percent change in SVI after VE (r = −0.45, 95% CI: −0.647– −0.207, P < 0.001 and r=0.465, 95% CI: 0.212−0.659, P < 0.001, respectively; Fig 5).

## Discussion

Nearly half of the patients in this study were classified as non-responders. In patients undergoing robot-assisted laparoscopic gynecological surgery in the modified head-down lithotomy position, the carotid FTc measured by Doppler ultrasound and ΔPPV_6–8_ obtained through the VT challenge reliably predicted fluid responsiveness. PPV is a reliable predictor after increasing VT to 8 mL/kg PBW; however, its predictive capacity at VT of 6 mL/kg PBW is inferior. Unlike PPV, SVV_6_ and SVV_8_ at low- or high-VT mechanical ventilation and absolute SVV change (ΔSVV_6–8_) obtained via VT challenge failed to predict fluid responsiveness. Except for FTc, PPV_8_ and ΔPPV_6-8_ determined the larger grey areas (almost half) for which fluid responsiveness cannot be reliably predicted; this may affect the clinical application.

Ultrasound imaging has been increasingly used in the perioperative period in recent years, and carotid ultrasonography is a potential tool for intraoperative fluid management. Compared with the aorta and peripheral arteries, the carotid artery is easily accessible and may be used to approximate the cardiac systolic flow time. The carotid FTc can be determined in a fast, simple, and non-invasive manner; thus, it is widely reported. Kim et al. [11] and Xu et al. [26] reported that FTc can predict fluid responsiveness in spontaneously breathing patients and provides the optimal cut-off value to predict fluid responsiveness; thus, FTc may be a substitute for dynamic indices relying on heart-lung interactions, including PPV. No manipulation of the VT setting or other additional invasive procedures are required in mechanically ventilated patients. The carotid FTc was also a good predictor of fluid responsiveness during low and high VT mechanical ventilation, indicating that variations in intra-thoracic pressure during respiration did not significantly affect carotid FTc [27,28]. Afterload or cardiac contractility can be affected by vasopressor infusion, thereby rendering the FTc values inaccurate. Therefore, changes in the carotid FTc may be a more useful predictor of fluid responsiveness than absolute FTc [22].

Our research aimed to overcome the limitations of conventional assessment methods including pneumoperitoneum, low ventilation volume, and body position-based variation in intra-thoracic pressure. Consistent with prior findings, this study showed that FTc can discriminate between fluid responders and non-responders. The threshold value of 356.5 ms (sensitivity, 84.6%; specificity, 69.2%) in this study was higher than those of other reports, possibly due to the cardiac output changes caused by the modified head-down lithotomy position, which unintentionally increased the false-negative rate; therefore, higher values were needed to identify affected patients. Furthermore, fluid responsiveness is defined as a ≥10%−15% increase in SVI, a range that allows cut-off values to vary due to a lack of uniformity; different researchers have chosen different values, and our study chose 10%. Our findings are consistent with those of Chen et al. [12], who found that FTc increased following fluid administration to the non-responsive group.

Intraoperative use of lung-protective ventilation strategies may reduce postoperative pulmonary complications; therefore, they are widely used in perioperative and critical situations [29]. However, the use of low VT ventilation is a common limitation of PPV and SVV use during controlled mechanical ventilation. PPV and SVV cannot reliably predict fluid responsiveness during low VT ventilation, regardless of the patient’s position (supine, prone, and Trendelenburg), possibly due to inadequate intra-thoracic pressure [7,15,16,30]. Similar to the findings of previous studies, pneumoperitoneum did not alter the ability of PPV to predict fluid responsiveness if VT was at least 8 mL/kg PBW [7,31]. Our optimal cut-off value for PPV was 8%, which is 1% higher than that reported by Jun et al. [7]; our results showed less sensitivity but more specificity, which may be due to differences in study design and patient population.

To improve dynamic indicators, functional hemodynamic tests combined with hemodynamic parameters, such as passive leg raising, end-expiratory occlusion, and mini-fluid challenge, may be used. The VT challenge test was proposed to improve the reliability of PPV and SVV measurement at low VT ventilation [30]. Myatra et al. [30] hypothesized that increasing VT from 6 to 8 mL/kg PBW increases the intra-thoracic pressure and magnitude of heart-lung interactions, thereby unmasking fluid responsiveness during low VT ventilation in responders, subsequently showing that the changes in PPV and SVV after the VT challenge identify true fluid responders, which has been recognized by numerous studies [7,15,16].

Our study also confirmed that changes in PPV allow surgeons to discriminate between fluid responders and non-responders; however, SVV at VT of 8 mL/kg PBW and the change in SVV after the VT challenge were poor predictors. According to Wajima et al. [32], SVV values must be estimated cautiously during pneumoperitoneum, and the ability of SVV to predict fluid responsiveness decreases after pneumoperitoneum is established. The changes in intra-thoracic pressure may be caused by intra-abdominal pressure alterations during pneumoperitoneum; however, this idea remains controversial and warrants further research [33]. SVV is affected by respiratory and posture factors, diminishing reliability in patients with poor lung compliance, excessively low VT values, or excessively high respiratory rates [34]. Furthermore, differences in operating principles and calculation methods of different devices (FloTrac/Vigileo, PiCCOplus, MostCare, etc.) may also contribute to inter-study differences in SVV values.

Our study has several limitations. Compared with the clinical gold standard of pulmonary artery catheters, our analysis of volume status and fluid responsiveness using uncalibrated pulse contouring of arterial waveforms may have been insufficient in hemodynamically stable patients, although the MostCare monitor estimated CO with a good level of agreement with echocardiographic measures [35]. Second, our study population included patients with gynecologic tumors; therefore, the findings might not apply to all patient populations. We did not consider the changes in peripheral vascular resistance caused by increased pressure of the arterial baroreceptor (carotid sinus) in the head-down position and the cerebral autoregulatory function activated by fluid redistribution.

In conclusion, FTc and ΔPPV_6–8_ are relatively reliable predictors of fluid responsiveness in patients undergoing robot-assisted laparoscopic gynecological surgery in the modified head-down lithotomy position with lung-protective ventilation. Moreover, since FTc is non-invasive, easily accessible, rapid, and reproducible, its measurement is particularly valuable for vessel volume assessment.

## Data Availability

All relevant data are within the manuscript and its Supporting Information files.

## Acknowledgements

None

## Supplemental Information

**Fig. S1.**
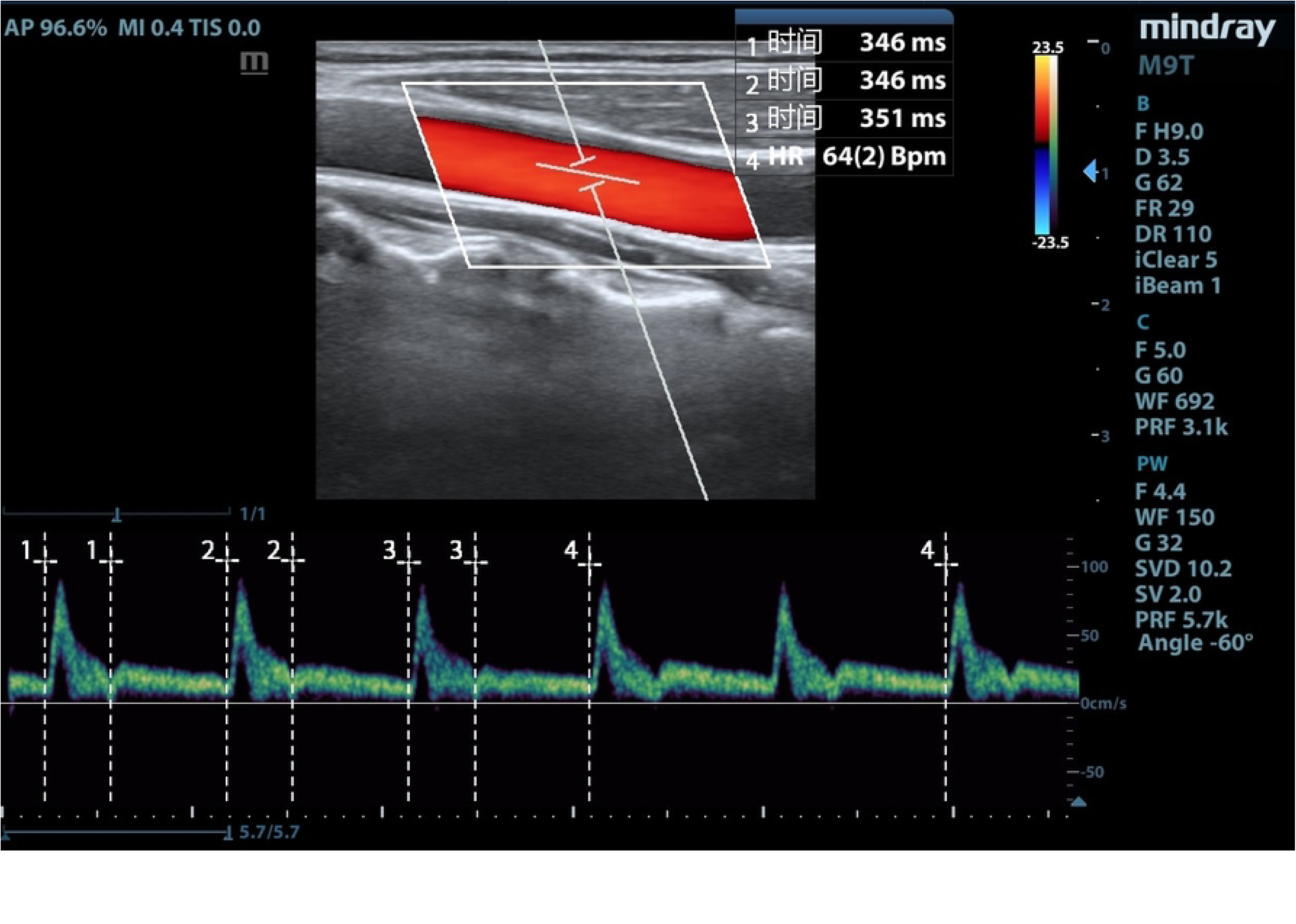
Carotid Doppler waveform. The numbers 1, 2, and 3 are flow time: beginning of the systolic upstroke to the dicrotic notch. Number 4 is the heart rate: the interval between the beginning of the two consecutive Doppler flow upstrokes.

**Fig. S2.**
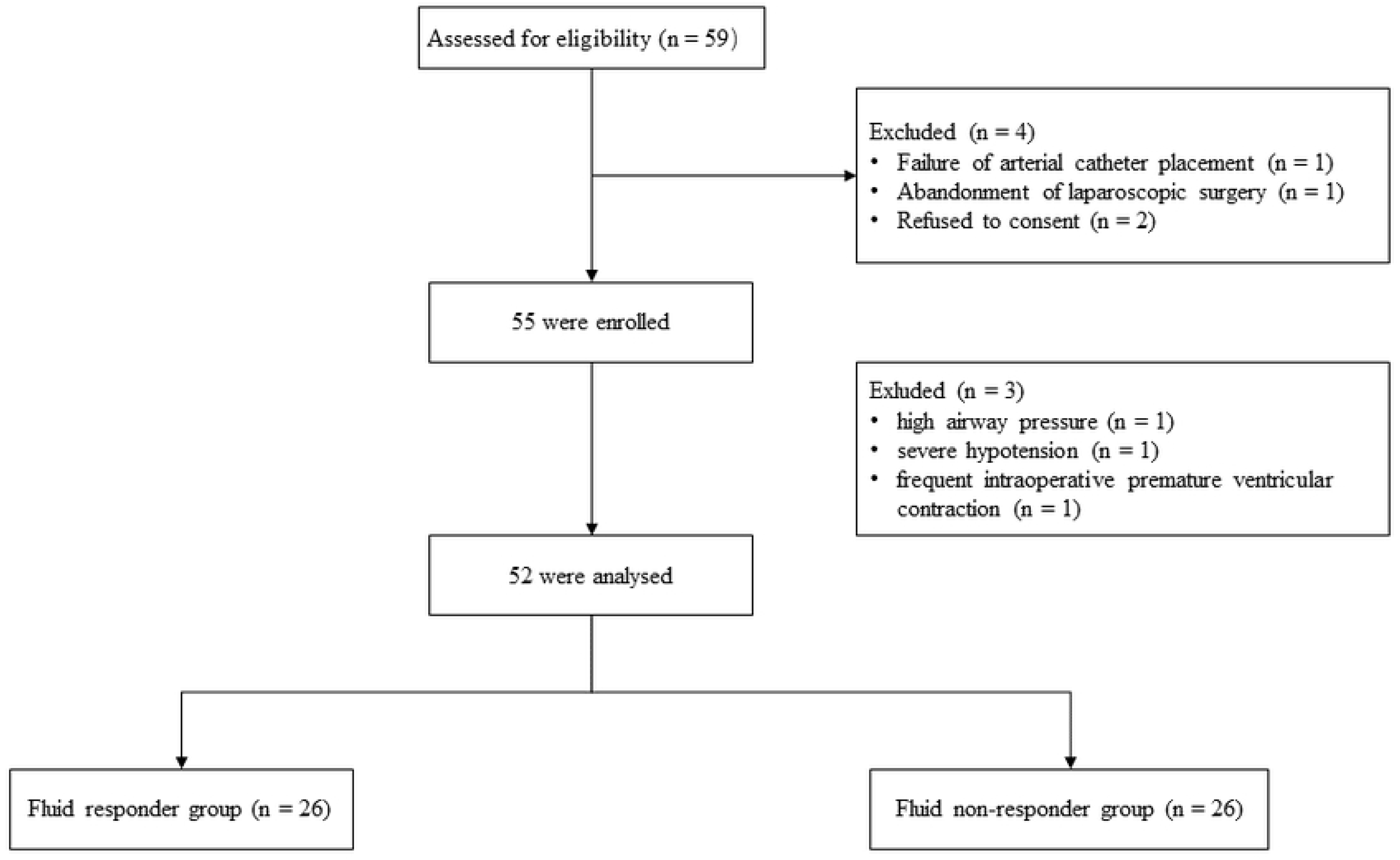
Flow chart of the number of patients enrolled in the study and included in the analyses.

## Notes

**Conflicts of Interest:** None

**Funding:** This work was supported by the Laboratory Open Fund of Chongqing University Cancer Hospital (2022); Chongqing medical scientific research project (Joint project of Chongqing Health Commission and Science and Technology Bureau 2023MSXM125)

### Competing Interest Statement

The authors have declared no competing interest.

### Funding Statement

The author(s) received no specific funding for this work

### Author Declarations

This study was approved by the Institutional Review Board of Chongqing University Cancer Hospital (approval number: CZLS2021041-A) and registered before patient enrollment on the Chinese Clinical Trial Register (CHiCTR2200060573)

